# In-home molecular testing of tongue swabs and sputum to inform household-level screening with diagnostic escalation strategies for tuberculosis contact investigation: a prospective cohort study in South Africa

**DOI:** 10.64898/2026.02.19.26346589

**Authors:** Andrew Medina-Marino, Sharon Olifant, Maria Pieruccini, Kuhle Fiphaza, Nondumiso Ngcelwane, Morten Ruhwald, Adam Penn-Nicholson, Bernard Fourie

## Abstract

**Background:** Household contact investigation for tuberculosis (TB) is limited by referral for clinic-based testing services. We evaluated the performance of in-home tongue swab (TS) testing among symptom-agnostic household contacts (HHC) to inform HCI screening strategies.

**Methods:** We conducted a prospective cohort study among HHC of TB patients in Eastern Cape, South Africa. In-home testing of sputum and TSs, with TSs pooled from up to three HHCs, was performed using Xpert Ultra on portable GeneExpert devices. Outcomes included diagnostic performance of TS testing relative to sputum and linkage-to-care outcomes.

**Findings:** Between June 2021 and October 2024, 909 HHC were enrolled; 99·1% provided s TS, 31·6% provided sputum. Overall sensitivity and specificity of TS testing was 61·9% (95% CI: 38·4%–81·9%) and 100% (98·9%–100%), respectively; sensitivity was 100% (47·8%–100%) for individually tested swabs. Among two-swab and three-swab pools where ≥1 individual was sputum positive, 55·6% (21·2%–86·3%) and 42·9% (9·9%–81·6%) tested positive, respectively; TS sensitivity declined with decreasing sputum Ultra semi-quantitative category. 27 of 439 (6·2%) households had an indictation of secondary TB; 13 (3·0%) by sputum and TS, 11 (2·5%) by sputum only, 3 (0·7%) by TS only. Sputum testing identified 29 HHC with TB (yield=3·2%); 25/29 (86·2%) linked to care (median 1 day [IQR 1–2]).

**Interpretation:** While in-home molecular testing of sputum supported rapid linkage-to-care, and TSs enabled near-universal testing of symptom-agnostic HHCs, efficiency gains through pooled TS testing must be balance against sensitivity trade-offs.

**Funding:** U.S. NIH; Australian Department of Foreign Affairs and Trade; UK Foreign, Commonwealth and Development Office

**RESEARCH IN CONTEXT:** *Evidence Before This Study:* Household contacts (HHCs) of people with TB are prioritized for active case-finding (ACF) strategies due to their increased risk of developing TB disease. Household contact investigation (HCI), a widely recommended ACF strategy, is constrained by attrition from referral-based cascades and sputum-based testing. We searched PubMed and Embase for studies published in English from January 1, 2010 to January 31, 2026, using combinations of the terms “tuberculosis” or “TB” with “household contact,” “contact tracing,” “contact investigation,” “screening,” “triage,” “in-home testing,” “molecular testing,” and “tongue swab.” We also reviewed references listed in relevant articles. There are limited data describing microbiological testing strategies targeting HHCs conducted outside clinic settings, and fewer still that explore the integration of HCI and in-home molecular TB testing. Tongue swabs have emerged as a promising non-invasive, non-sputum specimen type for molecular TB diagnosis. However, most tongue swab performance data have been generated in clinic-based or symptom-prompted populations, with a marked paucity of data generated in populations at high risk for asymptomatic or paucibacillary TB, including HHC. Before this study, published work exploring the use of tongue swabs within in-home TB testing strategies was limited to two papers, both from our group, which focused on acceptability, feasibility, and preliminary costing and modeling analyses. To date, no published studies have assessed the diagnostic performance of tongue swab–based molecular testing relative to sputum-based testing among HHC, the use of tongue swab specimens as part of in-home testing strategies, nor the implication of pooled tongue swab testing to inform household-level screening and diagnostic escalation strategies. In addition, evidence describing verified linkage to TB treatment services following in-home sputum molecular testing was limited to one pilot study paper.

*Added Value of This Study:* This study is the first to evaluate in-home molecular TB testing using tongue swab specimens, and to incorporate household-level pooling of tongue swabs from multiple household members as a primary screening strategy. Near-universal swab collection substantially expanded access to microbiological testing in a population with limited sputum production. Although pooled swab testing exhibited reduced sensitivity compared with individual-level sputum testing, stratified analyses of tongue swab tests by sputum Xpert Ultra semi-quantitative categories demonstrate that this reduction reflects a biological gradient associated with low mycobacterial burden. Importantly, pooled swab testing identified TB among contacts unable to produce sputum, increasing diagnostic yield beyond sputum-dependent approaches. The study also documents the increase in diagnostic yield when implementing a symptom-agnostic testing strategy among HHC, and rapid, verified linkage to clinic-based TB treatment services following in-home sputum testing.

*Implications of All the Available Evidence:* Collectively, the available evidence supports reframing TB household contact investigation from individual-level referral for clinic-based testing toward in-home testing models, including the use of household-level screening with diagnostic escalation. Near-universal, in-home collection of tongue swab specimens enables substantially broader microbiological assessment than sputum-dependent strategies and facilitates detection of TB among asymptomatic and sputum-scarce HHCs, individuals frequently missed by referral-based approaches for clinic-based sputum collection and testing. Our findings show that the reduced sensitivity associated with pooled tongue swab testing follows a predictable biological gradient related to mycobacterial burden rather than a technical failure of pooling. Pooled swab testing should therefore be understood as a household-level screening strategy within a sequential diagnostic algorithm, not a replacement for individual diagnosis. For TB programs, efficiency gains and expanded coverage achieved through pooling must be balance against sensitivity trade-offs. Household-level screening using pooled specimens can focus downstream referrals and may improve programmatic efficiency without requiring universal individual testing. Future research should evaluate optimized diagnostic algorithms that integrate pooled, non-sputum testing with diagnostic escalation, assess impact on linkage-to-care and prevention outcomes, and define the role of pooled testing within scalable, community-based TB case-finding strategies.

## Introduction

Tuberculosis (TB) household contact investigation (HCI) is a proactive strategy shown to increase early case detect, facilitate early treatment initiation and seeks to break chains of transmission. Systematic reviews and modelling studies estimate that a substantial proportion of incident TB arises among household contacts (HHC) of people with infectious TB, making contact investigation a high-yield strategy for early case detection and prevention.(1,2) Despite strong policy endorsement, programmatic implementation of HCI remains limited in many high-burden settings, with major gaps in coverage, diagnostic completion, and linkage-to-care.(3,4)

A central challenge is how TB services are accessed during contact investigation. Most programs rely on referral-dependent pathways in which TB patients are asked to refer their HHC to independently present for clinic-based screening, sputum collection and testing services, or contacts are screened for symptoms in the community and those with symptoms are referred for clinic-based sputum collection and testing. These approaches are poorly suited to the epidemiology of TB among HHC, in whom disease frequently exists along an asymptomatic and paucibacillary spectrum.(5) As a result, symptom-based screening is structurally misaligned with HHC risk, and sputum-dependent diagnostic strategies systematically constrain access, uptake, and case-finding yield.(6,7)

Tongue swabs (TS) have emerged as a promising, non-sputum-based sampling strategy to address these constraints by enabling near-universal specimen collection in community and household settings. Multiple studies demonstrate that swab-based specimens are acceptable, feasible, and preferred by individuals who are unable or unwilling to produce sputum.(8,9) Diagnostic accuracy studies have further shown that TS can detect TB using molecular platforms, although sensitivity varies and is generally lower than sputum, particularly among individuals with low bacillary load.(10,11)

Pooling of specimens represents an additional strategy to improve the efficiency and scalability of TB testing during HCI. Pooled testing has been evaluated using sputum as part of active case-finding activities, demonstrating potential benefits in terms of cartridge savings, staff time, and population-level impact when deployed strategically.(12,13) However, there is no published evidence on the use of non-sputum specimen pooling as part of household-level screening strategies for HCI, nor on how underlying bacterial burden shapes pooled test performance or how such approaches interface with downstream referral and care pathways.

To date, few studies have evaluated in-home TB testing models,(14,15) and none have explored the integration of near-universal non-sputum specimen collection, pooled molecular testing, or systematic assessment of linkage-to-care following microbiological confirmation. Addressing these gaps is increasingly important as TB programs seek strategies to expand and decentralize microbiological testing to support active case finding programs and identification of individuals with asymptomatic or paucibacillary TB.(8,9) This need is amplified for HHCs due to the high prevalence of asymptomatic and paucibacillary TB among this population, together with substantial attrition following clinic referral due to transport and other access barriers.(6,16)

To address these limitations and gaps, we conducted a prospective cohort study among HHC of people receiving treatment for drug-sensitive TB (DS-TB) in Eastern Cape, South Africa. We evaluated in-home collection and molecular testing of TS and sputum, using household-level pooling of TS specimens from different household members as the primary screening strategy. Our objectives were to assess the diagnostic performance of TS testing compared with sputum-based testing, including potential diagnostic trade-offs associated with pooling, and to evaluate linkage to clinic-based TB services following in-home microbiological confirmation.

## Methods

### Study Design and Setting

Between June 15, 2021, and October 29, 2024, we conducted a prospective cohort study among HHC of TB patients receiving treatment for DS-TB at 26 government health clinics in Buffalo City Metro (BCM) Health District, Eastern Cape Province, South Africa. In 2023, BCM had an estimated TB case notification of 900-999 per 100,000 population and HIV prevalence of 12·8%.(17,18)

### Participant Recruitment and Data Collection

Details regarding recruitment of TB index patients and their HHC have been described.(8,14) Briefly, individuals receiving treatment for DS-TB were identified and asked for permission to conduct a household visit. Following consent, a list of household members was recorded, and a home visit scheduled to occur within 3–4 days of recruitment. During household visits, staff confirmed household membership and screened individuals for eligibility (aged ≥18 years; not currently receiving TB treatment; did not complete TB treatment within past 6 months; written informed consent. Though HHC were screened for TB-related symptoms using the W4SS,(19) with cough of any duration, eligibility was independent of symptoms. Individuals who were ineligible or declined participation were offered a referral to a local clinic for further evaluation and services. Participant demographic, behavioral health and clinical data were collected by trained study staff using standardized electronic case report forms administered via REDCap,(20,21) and stored in U.S. HIPAA and South African POPI Act compliant servers.

### Specimen Collection and Testing

Contacts first provided three TS specimens. TS were collected by trained study staff using a Puritan HydraFlock swab [Puritan, Guilford, Maine, U.S.A.] rolled across the soft palate for 10 seconds, brushed across the tongue dorsum for 30 seconds, and rested on the tongue for an additional 10 seconds. Upon removal, the swab was placed into a tube containing 2 mL of PrimeStore® Molecular Transport Medium (PS-MTM) [Longhorn Vaccines & Diagnostics, San Antonio, Texas, U.S.A.]. Up to three TSs from different household members were included in a single tube of 2 mL PS-MTM regardless of pool size; households with more than three participants had additional pools tested in separate reactions. Swabs were incubated at room temperature for 15 minutes, after which 2 mL PS-MTM was loaded into a single GeneXpert [Cepheid, Sunnyvale, California, U.S.A.] MTB/RIF Ultra cartridge for immediate in-home testing. Although the study was designed around pooled TS testing, individual TS testing occurred when only a single HHC was present at the time of the visit, providing an opportunity to assess single swab performance. If a swab test was invalid or unsuccessful, the second collected swab was used for repeat testing. Untested swabs were biobanked for future use.

Sputum specimens were collected after that of TSs. Sputum were processed individually (i.e., not pooled) and tested in households according to manufacturer instructions and as previously described.(8,14) Multiple GeneXpert Omni (used June 2021–January 2022) or Xpert Edge (used June 2022–November 2024) instruments were operated concurrently as needed.

### Referrals for and Follow-up of Clinic-based TB Services

Details regarding referrals for clinic-based TB services have been described previously.(14,22) Briefly, contacts with a positive sputum result were referred immediately for clinic-based TB treatment. Symptomatic contacts and those unable to produce sputum but for whom a TS test was positive were referred for further clinical evaluation. In accordance with South African TB guidelines, those with negative test results (sputum or TS) were referred for clinic-based TPT services.(23) No transport support was provided, and study referrals did not receive preferential access to clinic services. Clinic presentation for TB services was ascertained through linkage with clinic registration and TB program records. Participants not identified through record reviews within 30 days were contacted by phone or follow-up household visit; those who could not be traced or whose self-reported attendance could not be verified were classified as lost to care.

### Statistical Analysis

Diagnostic performance of TS testing was evaluated against sputum-based molecular testing using Xpert MTB/RIF Ultra as the reference standard. The primary outcome was the diagnostic accuracy of TS testing compared with sputum Xpert Ultra, assessed using paired results irrespective of pooling strategy among participants who provided both specimen types with an available Xpert Ultra result. In this analysis, sputum Xpert Ultra results classified as trace were considered negative. Diagnostic accuracy measures (i.e., sensitivity and specificity) were estimated with exact 95% confidence intervals. Sample size was determined pragmatically based on available resources and the duration of the study period, with the aim of enrolling a cohort sufficient to estimate diagnostic performance with reasonable precision.

Secondary outcomes included diagnostic performance and implementation-relevant measures. Diagnostic performance was evaluated across pooling strategies, including single swabs, two-swab pools, and three-swab pools. Programmatic outcomes included the proportion of individuals with a positive sputum result that presented for clinic-based TB care and the time from referral-to-clinic presentation. Post-hoc exploratory analyses, conducted to aid interpretation of primary and secondary diagnostic performance findings, included stratification by sputum Xpert Ultra semi-quantitative categories (e.g., high, medium, low, very low, trace) to examine the relationship between underlying mycobacterial burden and TS test sensitivity.

Participant baseline characteristics were summarised using descriptive statistics, with categorical variables as frequencies with percentages and continuous variables presented as medians with interquartile ranges (IQR); where relevant, ranges (minimum–maximum) are also reported. Descriptive analyses were conducted using Stata [StataCorp, College Station, TX, USA]. Diagnostic accuracy estimates were calculated using MedCalc [MedCalc Software Ltd, Ostend, Belgium].

The Strengthening the Reporting of Observational Studies in Epidemiology (STROBE) guidelines were followed for reporting these data (Supplemental Table 1).(24)

### Ethical Considerations

Ethics approval was provided by the University of Pretoria (Ref. #57/2021) and University of Cape Town (Ref. #391/2021) research ethics committees. Permission was provided by the Eastern Cape Department of Health Provincial Research Committee (EC_202106_002). All participates provided written informed consent before participation.

## Results

### Participant Recruitment and Characteristics

From 439 households, 909 adult HHC were enrolled (Figure 1). Of those HHCs, 532 (58·5%) were asymptomatic (Table 1). The median age of enrolled HHC was 39 years (IQR: 28–55). Most participants were female (62·7%), black (71·5%), and unemployed (52·0%). Nearly half (47·5%) reported ever smoking, while most (65·5%) reported never or rarely drinking alcohol. Overall, 14·5% of participants self-reported living with HIV and 22·7% reported a prior history of TB, with a higher proportion of those experiencing TB-related symptoms reporting prior TB compared to those who were asymptomatic (29·4% vs. 18·0%).

**Figure 1:**
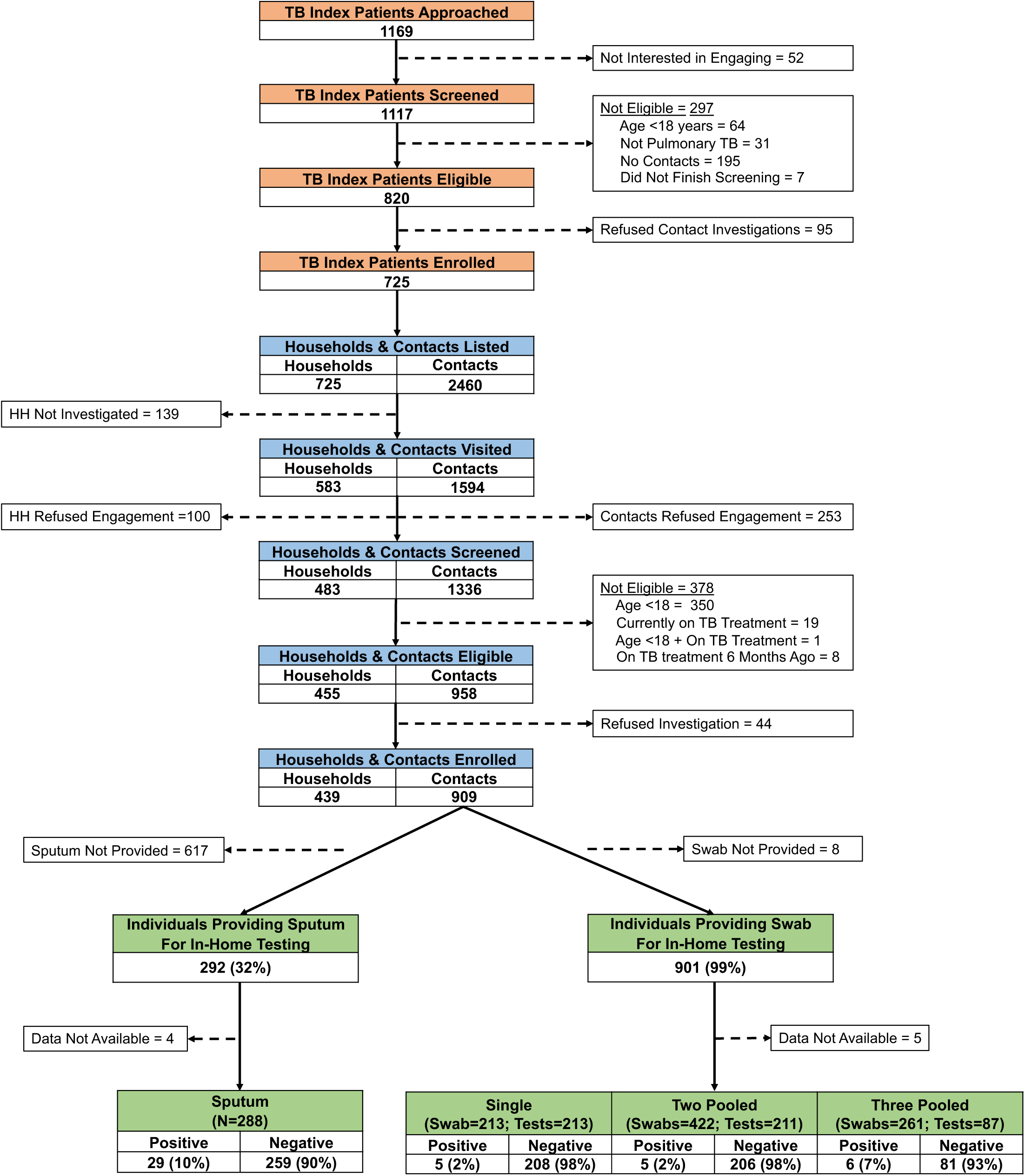
Study flow diagram.

**Figure 2:**
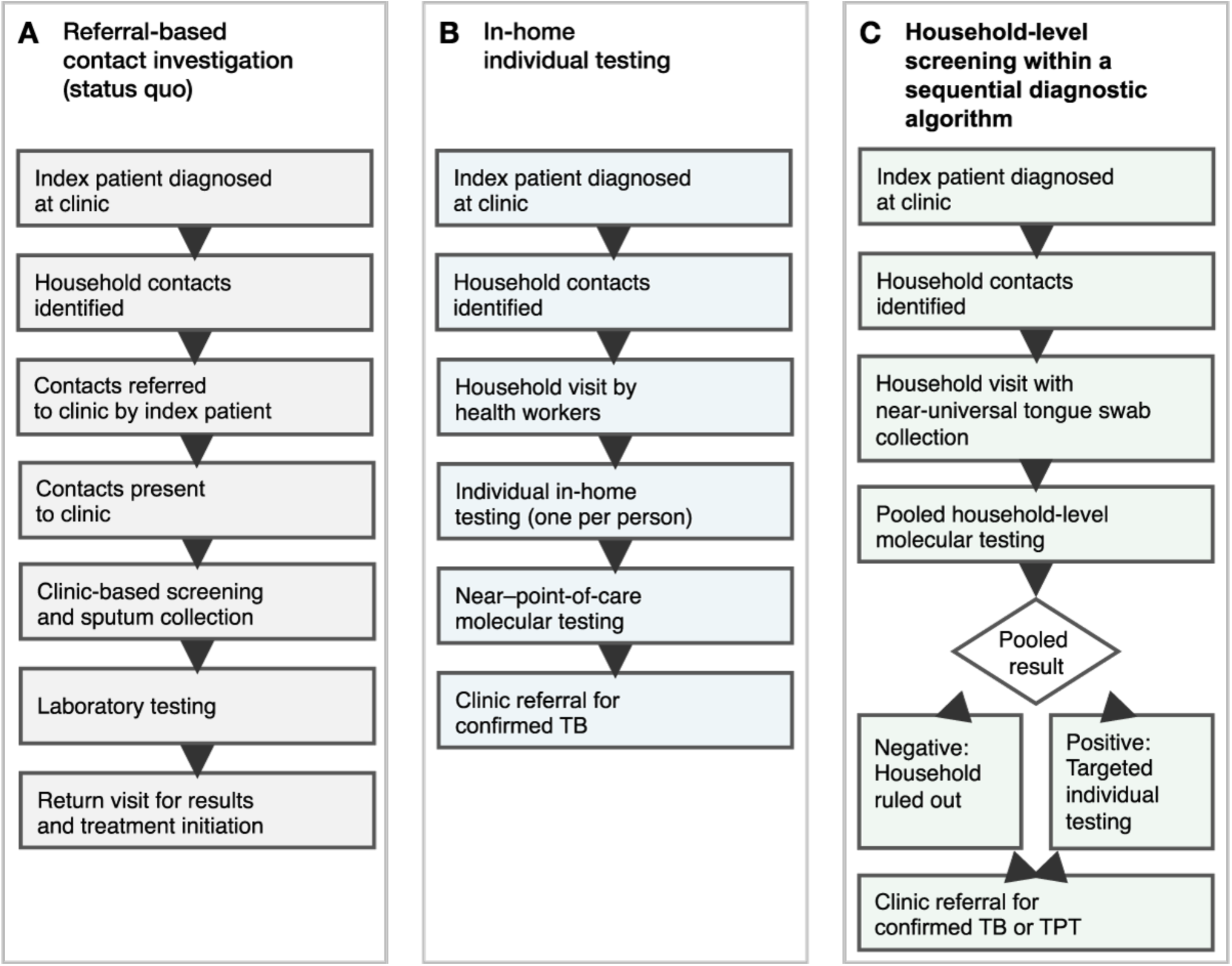
Status quo and conceptual pathways for tuberculosis contact investigation. **Panel A. Status quo pathways for TB contact investigation:** Panel A depicts commonly implemented contact investigation pathways following identification of an index TB patient. In many settings, household contacts are advised to attend clinics independently for evaluation or are symptom-screened during household visits and referred for facility-based testing. These approaches rely on sputum production and require multiple clinic visits for specimen submission, result receipt, and treatment initiation. At each step, substantial attrition occurs due to limited access, low uptake among asymptomatic contacts, inability to produce sputum, and delays inherent to referral-based diagnostic cascades. As a result, diagnostic coverage among exposed household members is incomplete, particularly for subclinical and paucibacillary TB. **Panel B. In-home individual testing:** Panel B illustrates an intermediate model in which molecular testing is conducted in the household on an individual basis. This approach reduces referral steps and improves uptake and completion of diagnostic evaluation compared with facility-based pathways. However, individual testing remains constrained by sputum dependence, requires one test per contact, and entails substantial cartridge use, staff time, and prolonged household visits, limiting scalability for households with multiple contacts or for broader community-based screening. Referrals for treatment or TPT are provided only to those with a sputum-based test result. **Panel C. Household-level screening within a sequential diagnostic algorithm:** Panel C presents the proposed household-level screening and individual-level diagnostic pathway informed by this study. Near-universal, non-sputum specimen collection using tongue swabs enables comprehensive household participation during a single visit. One or more pooled swab tests serve as an initial household-level screening step. Households with negative pooled results are ruled out without further testing. Positive pools trigger targeted individual molecular testing. Targeted referrals for either treatment or TPT can be made to all household contacts. This approach concentrates diagnostic resources where transmission is most likely, reduces cartridge use and staff time, minimizes diagnostic cascade attrition, and facilitates rapid linkage to care. When deployed strategically, with explicit consideration of sensitivity trade-offs, household-level screening offers a potentially scalable model for TB household contact investigation.

**Table 1:** Characteristics of the study population by tuberculosis phenotype.

| Characteristics | Total<br>(N=909) | Asymptomatic<br>household<br>contact with<br>TB<br>(n=13) | Symptomatic<br>household<br>contact with<br>TB<br>(n=16) | Asymptomatic<br>household<br>contact<br>without TB<br>(n=519) | Symptomatic<br>household<br>contact<br>without TB<br>(n=361) |
| --- | --- | --- | --- | --- | --- |
| **Median [IQR] age** | 39 [28, 55] | 37.5 [32, 44] | 39.0 [32, 44] | 39 [27, 55] | 40 [29, 56] |
| **Sex** |  |  |  |  |  |
| Female | 570 (62.7%) | 4 (30.8%) | 6 (37.5%) | 329 (63.4%) | 231 (64.0%) |
| Male | 338 (37.2%) | 9 (69.2%) | 10 (62.5%) | 190 (36.6%) | 129 (35.7%) |
| Missing | 1 (0.1%) | 0 (0%) | 0 (0%) | 0 (0%) | 1 (0.3%) |
| **Race** |  |  |  |  |  |
| Black | 650 (71.5%) | 6 (46.2%) | 11 (68.8%) | 379 (73.0%) | 254 (70.4%) |
| Colored | 250 (27.5%) | 6 (46.2%) | 5 (31.3%) | 135 (26.0%) | 104 (28.8%) |
| White | 2 (0.2%) | 1 (7.7%) | 0 (0%) | 0 (0%) | 1 (0.3%) |
| Indian | 4 (0.4%) | 0 (0%) | 0 (0%) | 4 (0.8%) | 0 (0%) |
| Other | 2 (0.2%) | 0 (0%) | 0 (0%) | 0 (0%) | 2 (0.6%) |
| Missing | 1 (0.1%) | 0 (0%) | 0 (0%) | 1 (0.2%) | 0 (0%) |
| **Employment Status** |  |  |  |  |  |
| Employed | 212 (23.3%) | 2 (15.4%) | 3 (18.8%) | 126 (24.3%) | 81 (22.4%) |
| Unemployed | 473 (52.0%) | 9 (69.2%) | 11 (68.8%) | 263 (50.7%) | 190 (52.6%) |
| Retired | 65 (7.2%) | 1 (7.7%) | 0 (0.0%) | 39 (7.5%) | 25 (6.9%) |
| Student | 40 (4.4%) | 0 (0%) | 0 (0%) | 31 (6.0%) | 9 (2.5%) |
| Sick/Disabled | 16 (1.8%) | 0 (0%) | 0 (0%) | 8 (1.5%) | 8 (2.2%) |
| Other | 102 (11.2%) | 1 (7.7%) | 2 (12.5%) | 51 (9.8%) | 48 (13.3%) |
| Missing | 1 (0.1%) | 0 (0%) | 0 (0%) | 1 (0.2%) | 0 (0%) |
| **History of smoking** |  |  |  |  |  |
| Never | 476 (52.4%) | 1 (7.7%) | 3 (18.8%) | 310 (59.7%) | 162 (44.9%) |
| Current | 360 (39.6%) | 12 (92.3%) | 13 (81.3%) | 175 (33.7%) | 160 (44.3%) |
| Former | 72 (7.9%) | 0 (0%) | 0 (0%) | 33 (6.4%) | 39 (10.8%) |
| Missing | 1 (0.1%) | 0 (0%) | 0 (0%) | 1 (0.2%) | 0 (0%) |
| **Alcohol Use** |  |  |  |  |  |
| Never | 439 (48.3%) | 9 (69.2%) | 8 (50.0%) | 258 (49.7%) | 164 (45.4%) |
| Rarely | 156 (17.2%) | 1 (7.7%) | 0 (0%) | 90 (17.3%) | 65 (18.0%) |
| Occasionally (1-3 days/week) | 267 (29.4%) | 3 (23.1%) | 6 (37.5%) | 145 (28.0%) | 113 (31.3%) |
| Frequently (4-6 days/week) | 36 (4.0%) | 0 (0%) | 1 (6.3%) | 21 (4.0%) | 14 (3.9%) |
| Daily (7 days/week) | 9 (1.0%) | 0 (0%) | 1 (6.3%) | 3 (0.6%) | 5 (1.4%) |
| Missing | 2 (0.2%) | 0 (0%) | 0 (0%) | 2 (0.4%) | 0 (0%) |
| **HIV Status** |  |  |  |  |  |
| Positive | 132 (14.5%) | 2 (15.4%) | 3 (18.8%) | 69 (13.3%) | 58 (16.1%) |
| Negative | 712 (78.3%) | 11 (84.6%) | 11 (68.8%) | 408 (78.6%) | 281 (77.8%) |
| Did not disclose | 64 (7.0%) | 0 (0%) | 2 (12.5%) | 41 (7.9%) | 22 (6.1%) |
| Missing | 1 (0.1%) | 0 (0%) | 0 (0%) | 1 (0.2%) | 0 (0%) |
| **Prior TB** |  |  |  |  |  |
| Never | 697 (76.7%) | 9 (69.2%) | 7 (43.8%) | 422 (81.3%) | 259 (71.7%) |
| Yes, <2 years ago | 53 (5.8%) | 2 (15.4%) | 4 (25.0%) | 19 (3.7%) | 28 (7.8%) |
| Yes, ≥2 years ago | 153 (16.9%) | 2 (15.4%) | 5 (31.3%) | 73 (14.1%) | 74 (20.5%) |
| **Times had TB in the Past** |  |  |  |  |  |
| 1 time | 171 (82.6%) | 3 (75.0%) | 9 (100.0%) | 76 (82.6%) | 83 (81.4%) |
| ≥2 times | 36 (17.4%) | 1 (25.0%) | 0 (0%) | 16 (17.4%) | 19 (18.6%) |
| **Cough** | 195 (21.5%) | - | 15 (93.8%) | - | 180 (49.9%) |
| **Fever** | 185 (20.4%) | - | 9 (56.3%) | - | 176 (48.8%) |
| **Night Sweats** | 153 (16.8%) | - | 10 (62.5%) | - | 143 (39.6%) |
| **Unexplained Weight Loss** | 136 (15.0%) | - | 12 (75.0%) | - | 124 (34.3%) |
| **Median (IQR) symptom count** | 1 [1,2] |  | 3 [2,4] |  | 1 [1,2] |
| **Ultra Semi-quantitative results** |  |  |  |  |  |
| Trace | 6 (20.7%) | 4 (30.8%) | 2 (12.5%) | - | - |
| Very Low | 4 (13·8%) | 1 (7·7%) | 3 (18·8%) | - | - |
| Low | 7 (24·1%) | 5 (38·5%) | 2 (12·5%) | - | - |
| Medium | 3 (10·3%) | 1 (7·7%) | 2 (12·5%) | - | - |
| High | 9 (31·0%) | 2 (15·4%) | 7 (43·8%) | - | - |
| **Presentation to the Clinic** |  |  |  |  |  |
| Yes | 25 (86·2%) | 9 (69·2%) | 16 (100%) | - | - |
| No/ Could Not Verify | 4 (13·8%) | 4 (30·8%) | 0 (0%) | - | - |
| **Time-to-Clinic Presentation** |  |  |  |  |  |
| Median, days [IQR] | 1 [1·0, 2·0] | 1 [1·0, 2·0] | 1 [0·5, 2·5] | - | - |
| Range, days (Min, Max) | 42 [0, 42] | 9 [0, 9] | 42 [0, 42] | - | - |
| **Clinic-based re-test Result** |  |  |  |  |  |
| Positive | 24 (96·0%) | 8 (88·9%) | 16 (100%) | - | - |
| Negative | 1 (8·0%) | 1 (11·1%) | 0 (0%) | - | - |
| **Initiated TB Treatment** |  |  |  |  |  |
| Yes | 24 (96·0%) | 8 (100%) | 16 (100%) | - | - |
| No | 0 (0%) | 0 (0%) | 0 (0%) | - | - |

### Specimen Collection and Test Results

Among HHCs, 99·1% (n=901) provided a TS specimen compared to 32.1% (n=292) that were sputum productive (Figure 1); a larger proportion of those reporting TB symptoms provided sputum compared to those who were asymptomatic (42·1% vs 25·0%; Table 1). Results were available for 288 of 292 (98·6%) sputum tested, of which 29/288 were positive for MTB (test positivity= 10·1%). Diagnostic yield under a symptom-restricted sputum collection and testing strategy was 1·8% (n=16/911), compared with 3·2% (n=29/911) under a symptom agnostic strategy (Table 1). Of those TS collected, 213/901 (23·6%) were tested as single swabs, 422 (46·8%) were tested in 211 pools of two swabs, and 261 (29·0%) were tested in 87 pools of three swabs (Figure 1). Among individually tested swabs, 5/213 (test positivity= 2·3%) yielded a positive result. Among pooled swab tests, 5/211 two-swab pools (test positivity= 2·4%) and 6/87 three-swab pools (test positivity= 6·9%) tested positive.

At a household level, 438/439 (99·8%) provided at least one TS specimen compared with 185/439 (42·1%) that provided at least one sputum (data not shown); all households from which at least one sputum was collected also provide at least one TS specimen. A total of 27 (6·2%) households had an indictation of secondary TB; 13 (3·0%) by both TS and sputum, 11 (2·5%) by sputum only, and 3 (0·7%) by swab only (data not shown).

### Linkage to Care Following Positive Sputum Result

All 29 individuals with a positive sputum result were referred for clinic-based TB services. Of those, 25/29 (86·2%) presented to a clinic within a median of 1 day (IQR: 1–2; range: 0–42 days); the four individual whose did not present to a clinic were asymptomatic (Table 2). In accordance with health department approvals, the 25 individuals who presented for clinic-based TB services were all re-tested, of which 24/25 (96·0%) re-tested positive; the single discordant result corresponded to an in-home sputum result classified as Xpert Ultra semi-quantitative trace. All 24 individuals with clinic-confirmed TB initiated treatment.

**Table 2:**
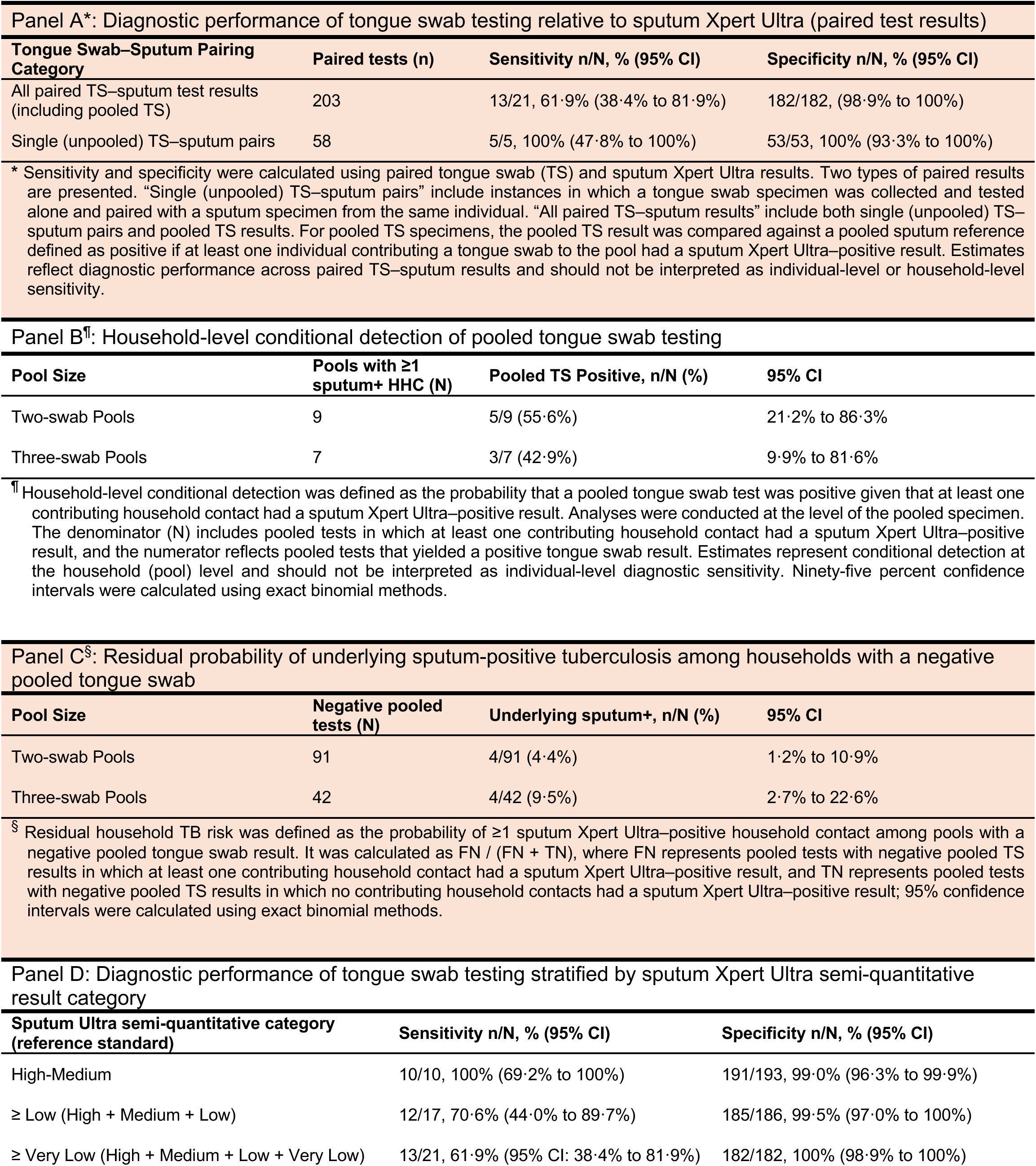
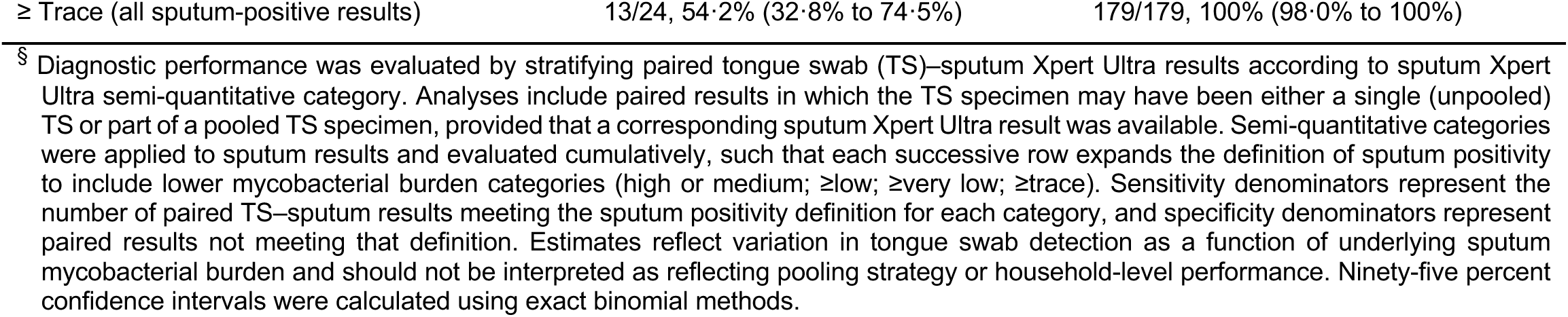
Evaluation of tongue swab testing strategies relative to sputum Xpert Ultra.

| Panel A*: Diagnostic performance of tongue swab testing relative to sputum Xpert Ultra (paired test results) |  |  |  |
| --- | --- | --- | --- |
| Tongue Swab–Sputum Pairing Category | Paired tests (n) | Sensitivity n/N, % (95% CI) | Specificity n/N, % (95% CI) |
| All paired TS–sputum test results (including pooled TS) | 203 | 13/21, 61·9% (38·4% to 81·9%) | 182/182, (98·9% to 100%) |
| Single (unpooled) TS–sputum pairs | 58 | 5/5, 100% (47·8% to 100%) | 53/53, 100% (93·3% to 100%) |
| * Sensitivity and specificity were calculated using paired tongue swab (TS) and sputum Xpert Ultra results. Two types of paired results are presented. “Single (unpooled) TS–sputum pairs” include instances in which a tongue swab specimen was collected and tested alone and paired with a sputum specimen from the same individual. “All paired TS–sputum results” include both single (unpooled) TS–sputum pairs and pooled TS results. For pooled TS specimens, the pooled TS result was compared against a pooled sputum reference defined as positive if at least one individual contributing a tongue swab to the pool had a sputum Xpert Ultra–positive result. Estimates reflect diagnostic performance across paired TS–sputum results and should not be interpreted as individual-level or household-level sensitivity. |  |  |  |
| Panel B<sup>†</sup>: Household-level conditional detection of pooled tongue swab testing |  |  |  |
| Pool Size | Pools with ≥1 sputum+ HHC (N) | Pooled TS Positive, n/N (%) | 95% CI |
| Two-swab Pools | 9 | 5/9 (55·6%) | 21·2% to 86·3% |
| Three-swab Pools | 7 | 3/7 (42·9%) | 9·9% to 81·6% |
| <sup>†</sup> Household-level conditional detection was defined as the probability that a pooled tongue swab test was positive given that at least one contributing household contact had a sputum Xpert Ultra–positive result. Analyses were conducted at the level of the pooled specimen. The denominator (N) includes pooled tests in which at least one contributing household contact had a sputum Xpert Ultra–positive result, and the numerator reflects pooled tests that yielded a positive tongue swab result. Estimates represent conditional detection at the household (pool) level and should not be interpreted as individual-level diagnostic sensitivity. Ninety-five percent confidence intervals were calculated using exact binomial methods. |  |  |  |
| Panel C<sup>§</sup>: Residual probability of underlying sputum-positive tuberculosis among households with a negative pooled tongue swab |  |  |  |
| Pool Size | Negative pooled tests (N) | Underlying sputum+, n/N (%) | 95% CI |
| Two-swab Pools | 91 | 4/91 (4·4%) | 1·2% to 10·9% |
| Three-swab Pools | 42 | 4/42 (9·5%) | 2·7% to 22·6% |
| <sup>§</sup> Residual household TB risk was defined as the probability of ≥1 sputum Xpert Ultra–positive household contact among pools with a negative pooled tongue swab result. It was calculated as FN / (FN + TN), where FN represents pooled tests with negative pooled TS results in which at least one contributing household contact had a sputum Xpert Ultra–positive result, and TN represents pooled tests with negative pooled TS results in which no contributing household contacts had a sputum Xpert Ultra–positive result; 95% confidence intervals were calculated using exact binomial methods. |  |  |  |
| Panel D: Diagnostic performance of tongue swab testing stratified by sputum Xpert Ultra semi-quantitative result category |  |  |  |
| Sputum Ultra semi-quantitative category (reference standard) | Sensitivity n/N, % (95% CI) |  | Specificity n/N, % (95% CI) |
| High-Medium | 10/10, 100% (69·2% to 100%) |  | 191/193, 99·0% (96·3% to 99·9%) |
| ≥ Low (High + Medium + Low) | 12/17, 70·6% (44·0% to 89·7%) |  | 185/186, 99·5% (97·0% to 100%) |
| ≥ Very Low (High + Medium + Low + Very Low) | 13/21, 61·9% (95% CI: 38·4% to 81·9%) |  | 182/182, 100% (98·9% to 100%) |

|  |  |  |
| --- | --- | --- |
| ≥ Trace (all sputum-positive results) | 13/24, 54·2% (32·8% to 74·5%) | 179/179, 100% (98·0% to 100%) |

<sup>§</sup> Diagnostic performance was evaluated by stratifying paired tongue swab (TS)–sputum Xpert Ultra results according to sputum Xpert Ultra semi-quantitative category. Analyses include paired results in which the TS specimen may have been either a single (unpooled) TS or part of a pooled TS specimen, provided that a corresponding sputum Xpert Ultra result was available. Semi-quantitative categories were applied to sputum results and evaluated cumulatively, such that each successive row expands the definition of sputum positivity to include lower mycobacterial burden categories (high or medium; ≥low; ≥very low; ≥trace). Sensitivity denominators represent the number of paired TS–sputum results meeting the sputum positivity definition for each category, and specificity denominators represent paired results not meeting that definition. Estimates reflect variation in tongue swab detection as a function of underlying sputum mycobacterial burden and should not be interpreted as reflecting pooling strategy or household-level performance. Ninety-five percent confidence intervals were calculated using exact binomial methods.

### Diagnostic Performance of Tongue Swab Testing

Diagnostic performance of TS testing compared with sputum-based molecular testing is shown in Table 2. When all TS tests were considered together regardless of pooling (203 paired tests), overall diagnostic performance relative to sputum Xpert Ultra demonstrated a sensitivity of 61·9% (95% CI, 38·4–81·9%) and a specificity of 100% (98·9–100%) (Panel A). Among individually tested TSs paired with sputum results (58 paired tests), sensitivity was 100% (47·8–100%) and specificity was 100% (93·3–100%).

For pooled TS testing, the unit of analysis was the pooled specimen rather than the individual. Among pooled TS tests in which at least one contributing HHC had a sputum positive result, 55·6% (21·2–86·3%) of two-swab pools and 42·9% (9·9–81·6%) of three-swab pools tested positive (Panel B). These estimates reflect conditional detection of TB at the household level (i.e., the probability that a pooled test is positive given at least one sputum-positive HHC). Among households with a negative pooled TS result, the residual probability of an underlying individual with sputum positive TB was 4·4% (4/91; 95% CI, 1·2–10·9%) for two-swab pools and 9·5% (4/42; 2·7–22·6%) for three-swab pools (Panel C). Pooled swabs were more likely to test positive when at least one contributing individual provided a sputum with a Xpert Ultra semi-quantitative result of high or medium, whereas pooled swabs were more frequently negative when sputum contributed by individuals all tested low, very low, or trace (Supplemental Figure 1).

To examine how detection by TS testing varied with underlying mycobacterial burden, diagnostic performance was stratified by sputum Xpert Ultra semi-quantitative categories (Table 2, Panel D) using paired TS–sputum results, irrespective of pooling strategy. When sputum positivity was defined as Xpert Ultra semi-quantitative high or medium, TS testing demonstrated 100% sensitivity (69·2–100%) and 99·0% specificity (96·3–99·9%). Expanding the positive reference category to include low semi-quantitative sputum results reduced sensitivity to 70·6% (44·0–89·7%). Further declines in sensitivity were observed as very low (61·9%; 38·4–81·9%) and trace (54·2%; 32·8–74·5%) results were included; specificity remained >99% across all definitions. These findings indicate a graded decline in TS sensitivity corresponding to decreasing sputum mycobacterial burden.

## Discussion

In this study of in-home TB testing of HHC using sputum and TS, we report three key findings. First, TS collection and testing substantially expanded diagnostic coverage in a population with a high prevalence of asymptomatic and paucibacillary disease, for whom sputum production is limited. Second, reduced sensitivity of TS testing was associated with lower underlying mycobacterial burden as reflected by sputum Xpert Ultra semi-quantitative results, indicating a biological gradient rather than a technical limitation of pooling per se. Third, in-home molecular testing supported rapid linkage to care, with most individuals with microbiologically confirmed TB presenting promptly for clinic-based services and initiating treatment. Together, these findings suggest that in-home testing of pooled, swab-based specimens can shift TB contact investigation from individual-level referral toward household-level screening with diagsnotic escalation, while highlighting the need for programmatic deployment that explicitly accounts for reduced sensitivity among individuals with low mycobacterial burden.

In our cohort, fewer than one-third of contacts were sputum productive, whereas nearly all provided a TS. This demonstrates how sputum-dependent diagnostic strategies constrain access, uptake, and case-finding yield during community-based HCI.(7,25) Implementation of near-universal specimen collection addresses the substantial proportion of TB that exists along an asymptomatic and paucibacillary spectrum, for which symptom-based screening and sputum-dependent diagnostics are poorly suited.(1,5) Connected to this, in-home testing directly addresses well-documented attrition in referral-based diagnostic cascades, where substantial losses occur when contacts are required to travel to clinics for symptom screening and sputum submission.(3,16) Together, this study supports swab-based, in-home testing as a pragmatic approach to closing persistent diagnostic access gaps in HCI and aligns with emerging evidence that TSs can equitably expand access to TB molecular testing services.(8,14)

Our finding of a reduced sensitivity of TS, particularly when TS pools include individuals with lower sputum bacterial burden, is consistent with prior studies.(11,10,26) Importantly, among pooled TS tests, unpaired positive pools demonstrate microbiological detection among individuals unable to produce sputum who would otherwise have remained untested and unidentified using sputum-based testing alone. This highlights the potential for swab-based testing to increase diagnostic yield beyond sputum-restricted strategies. Emerging evidence suggests that pooling approaches may be enhanced by incorporating bacteriologically richer specimens, such as sputum-swabs, which have demonstrated higher sensitivity than TS across multiple molecular platforms, including those designed for swab-based testing and decentralized use.(25,27) Together, these data suggest that household-level pooling strategies can likely be optimized by prioritizing the inclusion of bacteriologically richer sputum-swabs from those who are sputum productive with TS specimens from those who are sputum scarce. Such an approach can thus maximize achievable sensitivity while retaining the efficiency advantages of pooled swabs.

From a primary care and programmatic delivery perspective, pooled swab-based testing is best understood not as a replacement for individual diagnosis, but as a household-level screening strategy within a sequential diagnostic algorithm (Figure 1). By enabling a single pooled molecular test to serve as an initial rule-out or rule-in step at the household level, this approach simplifies decision-making in settings where multiple HHC require timely screening and diagnostic evaluation. When pooled results are positive, targeted individual testing concentrates diagnostic resources where transmission risk is highest. When pooled results are negative, no further diagnostic escalation is required within the household, conserving cartridges, staff time, and visit duration. In this way, household-level screening distinguishes households requiring further diagnostic evaluation for TB disease from those that can be prioritized for TPT, sharpening downstream referral rather than eliminating it.(28) Consistent with synthesis, modeling, and cost-effectiveness analyses, pooled testing can confer population-level benefits when deployed strategically, provided that sensitivity trade-offs are explicitly incorporated into program design and downstream diagnostic algorithms.(13,29) Within this framework, the programmatic value of household-level screening is realized through its downstream effects on care pathways, particularly improved timeliness and completeness of the diagnostic cascade for HHC and reduced attrition between community-based screening and clinic-based care.

Beyond diagnostic performance, our findings demonstrate that in-home molecular testing can support rapid linkage to TB treatment services, with most individuals with microbiologically confirmed TB presenting for care within days of referral. Importantly, among the small number of individuals who did not present for clinic-based services despite a positive sputum result, all were asymptomatic at the time of diagnosis. This highlights a critical implementation consideration as access to microbiological testing expands beyond symptom-based pathways. Prior work has shown that substantial attrition when contacts are referred for clinic-based evaluation, particularly when individuals do not perceive themselves to be ill.(3,16) As non-sputum–based testing approaches increasingly enable microbiological confirmation of TB among asymptomatic or minimally symptomatic contacts, linkage-to-care efforts must extend beyond diagnostic access and include tailored counseling, risk communication, and referral support that explicitly address perceived wellness and the benefits of early treatment initiation. Without such adaptations, expanded diagnostic reach may not fully translate into timely treatment or prevention uptake. These findings underscore that the population-level impact of decentralized, non-sputum–based TB testing will depend not only on diagnostic performance, but also on deliberate investments in linkage strategies designed for earlier and less symptomatic disease.

This study represents the first evaluation of integrating in-home molecular testing of HHC using single and pooled TS specimens, and documentation of verified linkage-to-care outcomes. Near-universal testing of all HHC using TSs strengthens the relevance of our approach to real-world settings where sputum scarcity and asymptomatic disease are common. However, diagnostic accuracy estimates were constrained by the limited number of paired sputum–TS comparisons and the small number of microbiologically confirmed TB outcomes inherent to the study’s pilot design, resulting in wide confidence intervals for sensitivity. Although inclusion of sputum-scarce contacts limited precision, it enhances external validity compared with studies restricted to sputum-producing individuals. As a single-arm cohort study, causal inference regarding effectiveness relative to referral-based diagnostic pathways is limited. TS testing was performed using molecular platforms and workflows adapted from sputum-based use rather than systems specifically optimized for swab specimens or decentralized in-home testing. Performance and operational efficiency may therefore differ as purpose-built swab platforms become more widely available. Finally, though our study did not detect drug-resistant TB in sputum or TS specimens, new swab-based platforms must support linkage to appropriate treatment regimens. Larger comparative and implementation studies are needed to more precisely define the role of household-level screening strategies enabled by in-home testing.

Pooled, swab-based testing, if appropriately optimized, may enable a shift from individual-level referral for clinic-based testing toward household-level screening with targeted individual diagnostic escalation, allowing diagnostic resources to be used more efficiently while maintaining timely identification of individuals requiring further evaluation or care. As testing of non-sputum specimens expands and decentralized molecular platforms continue to evolve, household-level screening and in-home diagnostic testing offers a promising framework to strengthen TB contact investigation, improve completion of diagnostic cascades, and support more targeted referral and delivery of TB preventive and treatment services.

## Data Availability

All data produced in the present study are available upon reasonable request to the authors

## CONTRIBUTORS

A.M.-M. conceptualized the study, developed the methodology, conducted formal analyses, drafted the original manuscript, supervised the study, and acquired funding. S.O. contributed to study conceptualization and methodology and provided critical review and editing of the manuscript. M.P. curated the data, conducted formal analyses, validated findings, prepared visualizations, and contributed to drafting the manuscript. K.F. conducted investigations, curated data, prepared visualizations, supported project administration, and reviewed and edited the manuscript. N.N. contributed resources, project administration, and manuscript review and editing. M.R. contributed resources and manuscript review and editing. A.P.-N. contributed to study conceptualization, manuscript review and editing, and funding acquisition. B.F. contributed to study conceptualization, methodology, formal analysis, supervision, and funding acquisition.

All authors had full access to the data, verified the integrity and accuracy of the data and analyses, agreed with the interpretation of the findings, and reviewed and approved the final manuscript. The corresponding author had final responsibility for the decision to submit for publication.

## DATA SHARING

De-identified study data are available from the corresponding author upon reasonable request.

## ACKNOWLEDGEMENTS

We thank the Eastern Cape Provincial Department of Health and the Buffalo City Metropolitan Health Department for their support and permission to conduct this study. We thank Samuel G. Schumacher for conceptual support and Grant Theron for critical review and feedback on previous drafts of this manuscript. We further thank Dana Bezuidenhout, Charl Bezuidenhout, and Nkosi Sibanda for their key contributions to project planning, implementation, field staff management, and data management. We also thank the field staff for their commitment and the study participants for their willingness to take part in this research. We acknowledge Cepheid for training, technical support, and the provision of Omni and Edge devices and Xpert MTB/RIF Ultra cartridges.

## DISCLAIMER

The content is solely the responsibility of the authors and does not necessarily represent the official views of the U.S. National Institutes of Health, the Australian Department of Foreign Affairs and Trade, or the UK Foreign, Commonwealth and Development Office.

## USE OF ARTIFICIAL INTELLIGENCE

During manuscript preparation, the authors used Chat GPT 5.2 to improve the readability of the manuscript. After using this tool, the authors reviewed and edited the content as needed; they take full responsibility for the content of the publication

---

**Supplemental Figure 1:**
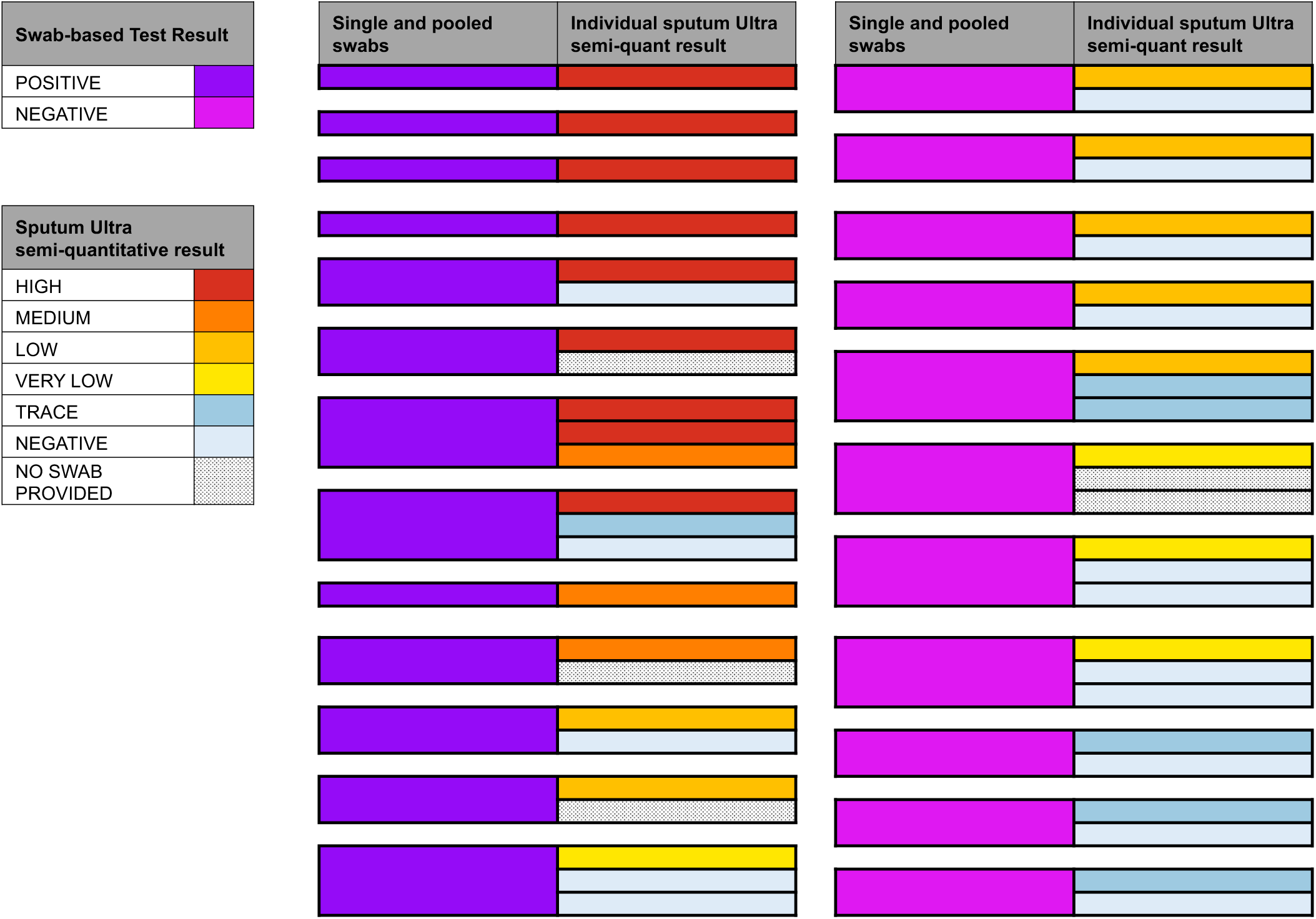
Relationship between sputum Xpert Ultra semi-quantitative test results and paired individual or pooled tongue swab test results.

